# Heterogeneous changes in mobility in response to the SARS-CoV-2 Omicron BA.2 outbreak in Shanghai

**DOI:** 10.1101/2023.05.12.23289890

**Authors:** Juanjuan Zhang, Suoyi Tan, Cheng Peng, Xiangyanyu Xu, Mengning Wang, Wanying Lu, Yanpeng Wu, Bin Sai, Mengsi Cai, Allisandra G. Kummer, Zhiyuan Chen, Junyi Zou, Wenxin Li, Wen Zheng, Yuxia Liang, Yuchen Zhao, Alessandro Vespignani, Marco Ajelli, Xin Lu, Hongjie Yu

## Abstract

The coronavirus disease 2019 (COVID-19) pandemic and the measures taken by authorities to control its spread had altered human behavior and mobility patterns in an unprecedented way. However, it remains unclear whether the population response to a COVID-19 outbreak varies within a city or among demographic groups. Here we utilized passively recorded cellular signaling data at a spatial resolution of 1km × 1km for over 5 million users and epidemiological surveillance data collected during the SARS-CoV-2 Omicron BA.2 outbreak from February to June 2022 in Shanghai, China, to investigate the heterogeneous response of different segments of the population at the within-city level and examine its relationship with the actual risk of infection. Changes in behavior were spatially heterogenous within the city and population groups, and associated with both the infection incidence and adopted interventions. We also found that males and individuals aged 30-59 years old traveled more frequently, traveled longer distances, and their communities were more connected; the same groups were also associated with the highest SARS-CoV-2 incidence. Our results highlight the heterogeneous behavioral change of the Shanghai population to the SARS-CoV-2 Omicron BA.2 outbreak and the its effect on the heterogenous spread of COVID-19, both spatially and demographically. These findings could be instrumental for the design of targeted interventions for the control and mitigation of future outbreaks of COVID-19 and, more broadly, of respiratory pathogens.

**Significance Statement:** Our study utilized passively recorded cellular signaling data and epidemiological surveillance data to investigate the changes human mobility to a COVID-19 outbreak at an unprecedented within-city level and examine its relationship with the actual risk of infection. Our findings highlight the heterogeneous behavioral change of the Shanghai population to the 2022 SARS-CoV-2 Omicron BA.2 outbreak and its heterogenous effect on the SARS-CoV-2 spread, both spatially and demographically. The implications of our findings could be instrumental to inform spatially targeted interventions at the within-city scale to mitigate possible new surges of COVID-19 cases as well as fostering preparedness for future respiratory infections disease outbreaks.

## Introduction

Following the initial COVID-19 wave in early 2020, mainland China adopted stringent measures, often referred to as the “zero-COVID” strategy, to curb COVID-19 outbreaks(1). This approach effectively minimized SARS-CoV-2 transmission in China until the emergence of the Omicron variant in late 2021(2). Subsequently, several Omicron outbreaks occurred, with a significant outbreak in Shanghai, identified in March 2022, accounting for over 600,000 confirmed infections(3). Comprehensive PCR testing, citywide lockdowns, and additional measures to restrict interpersonal interactions were implemented, ultimately containing the outbreak by June 2022. China eventually abandoned the “zero-COVID” policy six months later(4).

Human mobility patterns, ranging from international travel to daily commuting, significantly influence the spread of infectious diseases due to the nature of interpersonal interactions(5-10). Recent years have seen an exponential growth in geolocated datasets that provide unprecedented levels of detail to quantify human mobility(10-19). In particular, data collected from mobile devices has extensively been used in the early phase of the COVID-19 pandemic to investigate transmission dynamics, estimate changes in contact patterns as a result of public health interventions, and forecast epidemic spread(11, 13, 18, 19). However, limitations in the epidemiological and mobility data analyzed (e.g., varying COVID-19 reporting rates by age, incomplete demographic information for individual travel trajectories) have left several key questions regarding the relationships between epidemic spread, implemented interventions, and human behavior and mobility unanswered. In particular, it remains unclear whether population responses to a COVID-19 outbreak, as measured by travel frequency, distance traveled, and population connectivity, vary within a city (e.g., by district area) or among demographic groups (e.g., by age and sex).

To address these knowledge gaps, we utilized passively recorded Cellular Signaling Data (CSD) from over 5 million users (approximately 20% of Shanghai’s population) and epidemiological surveillance data collected during the SARS-CoV-2 Omicron outbreak in Shanghai. The exceptional scale and resolution of the human mobility data enabled us to analyze micro-level changes in mobility within the city and among different population groups (e.g., age and sex). Additionally, the repeated city-wide PCR screenings provided an opportunity to examine the association between these behavioral shifts and high-quality epidemiological data in the unique context of Shanghai’s 2022 Omicron outbreak.

## Results

### Omicron outbreak in Shanghai and public health response

In early March 2022, Shanghai experienced a significant outbreak of the SARS-CoV-2 Omicron variant, which rapidly spread among its 25 million residents. Throughout the outbreak, authorities conducted multiple mass PCR screenings; by the end of the outbreak on June 30, 2022, they had identified a total of 627,132 SARS-CoV-2 infections (see Fig. 1a). During the outbreak’s initial phase, authorities implemented grid management and partial lockdowns at the subdistrict level. On March 28, eastern Shanghai, consisting of subdistricts east of the Huangpu River (see SI Appendix, Fig. S1a), entered a population-wide lockdown, followed by a citywide lockdown for the rest of Shanghai on April 1. The citywide lockdown was lifted entirely on June 1, 2022, when the daily number of newly reported infections dropped to 10. Further information on the public health response can be found in the Methods, SI Appendix, Fig. S2, and SI Appendix, Table S2.

**Figure 1.**
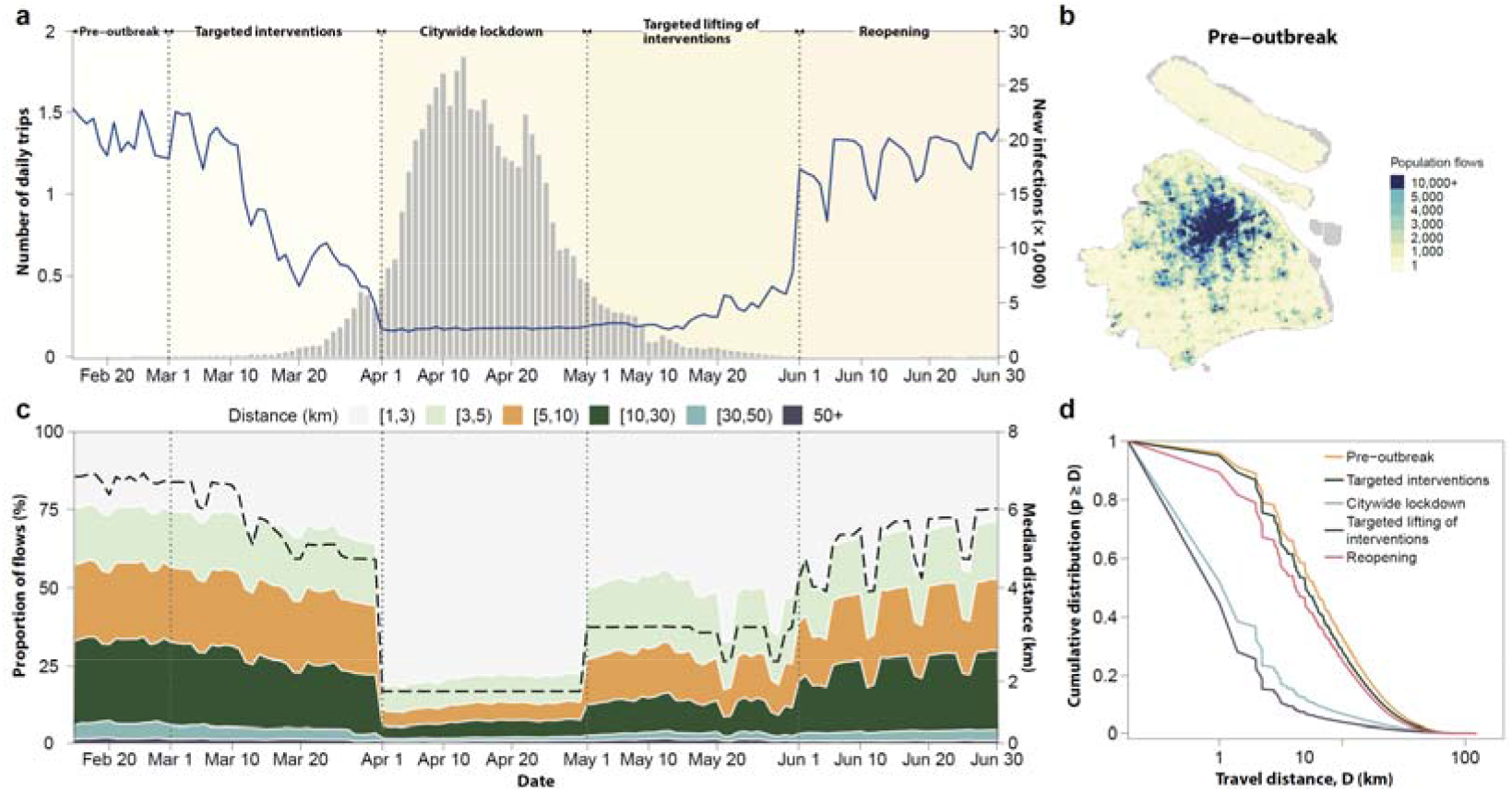
Changes in population flows and travel distance in Shanghai. **a**. Changes in number of daily trips and number of new infections reported from February 15 to June 30, 2022. Grey bars represent the daily reported infections. **b**. The geographic distribution of population trips during the pre-outbreak phase. The color intensity represents the number of daily trips occurred in each cell. **c**. The proportion of daily trips by different distances travelled (filled colors) and median distance of daily trips (dotted line) from February 15 to June 30, 2022. **d**. The cumulative probability distribution against distance (log) of daily trips across all five phases, where *p* is defined as the probability of traveling between locations at a certain distance. Each line represents the probability distribution per phase.

### Changes in frequency of travel, distance traveled, and mobility network community structure over the course of the outbreak

We quantified spontaneous and intervention-induced behavioral changes of the Shanghai population in terms of their daily mobility patterns based on CSD. During the study period (see SI Appendix, Fig. S1e), we analyzed an average of 5.04 million users accounting for 27% of all mobile phone users in Shanghai (20% of the total population). We estimated aggregated mobility flows, defined as the number of trips between two locations where a user spends at least 30 minutes, at a spatial resolution of 1km × 1km (see SI Appendix, Fig. S1d). This was done using a grid comprising 7,355 cells that covered the entire city of Shanghai, including all of its 16 districts and 216 subdistricts (see SI Appendix, Fig. S1a). The geographical distance between the cell centroids it is assumed to estimate the travel distance. Subsequently, we employed the Infomap method(20) to identify community structures within the mobility networks. Further details can be found in the Methods section and SI Appendix Section 1.

### Pre-outbreak Phase

During the two weeks before the Omicron outbreak began, we estimated an average of 1.36 trips per individual per day, corresponding to a total of 7.03 million trips per day (see Fig. 1a). Approximately, 33.4% of the grids in the central urban areas accounted for 80% of the total mobility in Shanghai (see Fig. 1b). The median distance traveled was 6.04 km; trips within 10 km accounted for 66.7% of all trips (see Fig. 1c and d). We identified 22 total communities, with a sizable core community (∼65.3% of Shanghai’s land area) at the city’s center, surrounded by peripheral communities outside the Shanghai metropolitan area (see Fig. 2a and SI Appendix, Fig. 3a and f).

**Figure 2.**
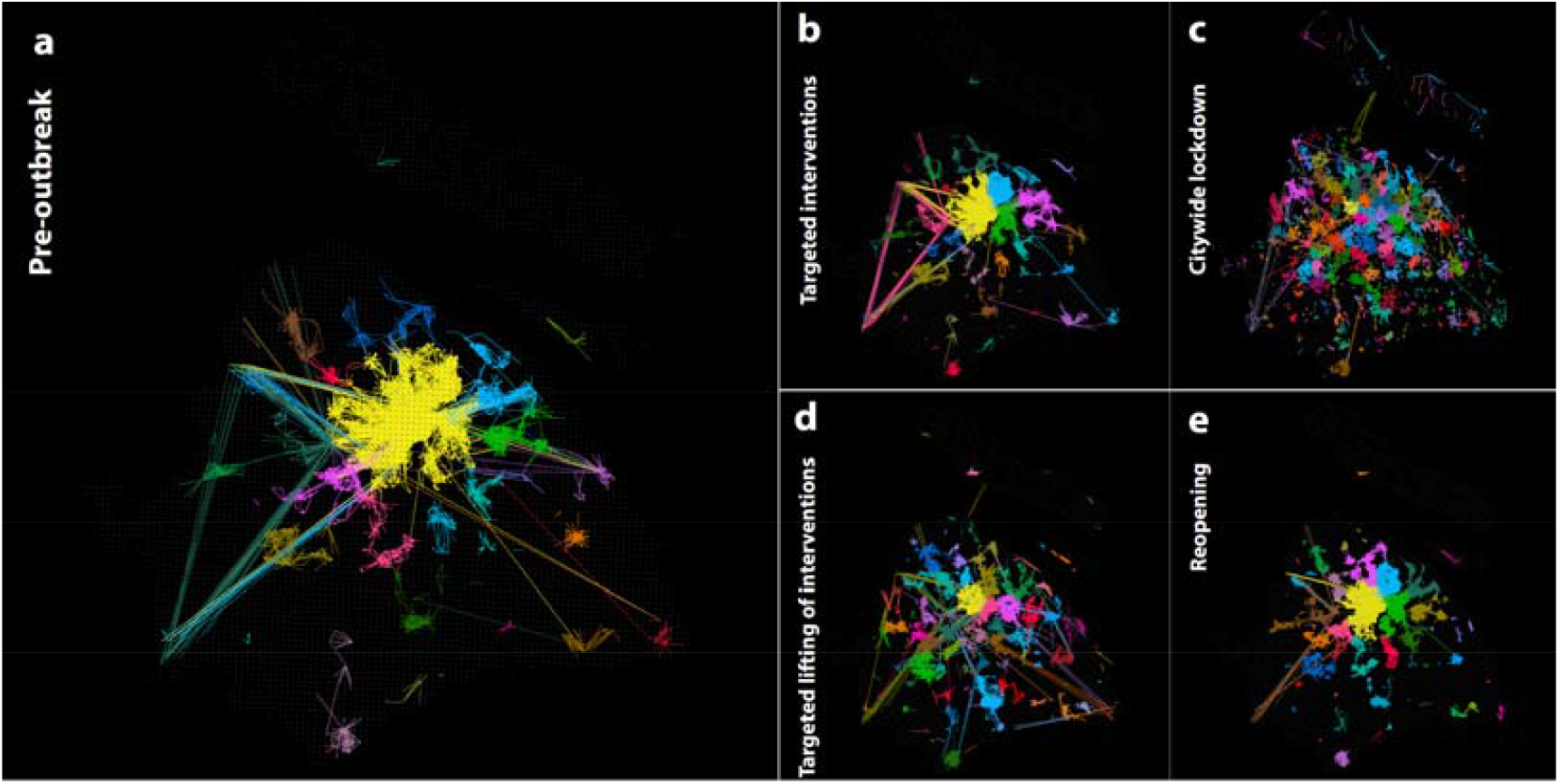
The network structural changes during each phase. **a-e**. The community structure of pre-outbreak, targeted interventions, citywide lockdown, targeted lifting of interventions, and reopening phases, respectively. The mobility network is visualized with the top 10,000 edges sorted by weight in descending order. The color of the edges illustrates the community partitions of the grid.

**Figure 3.**
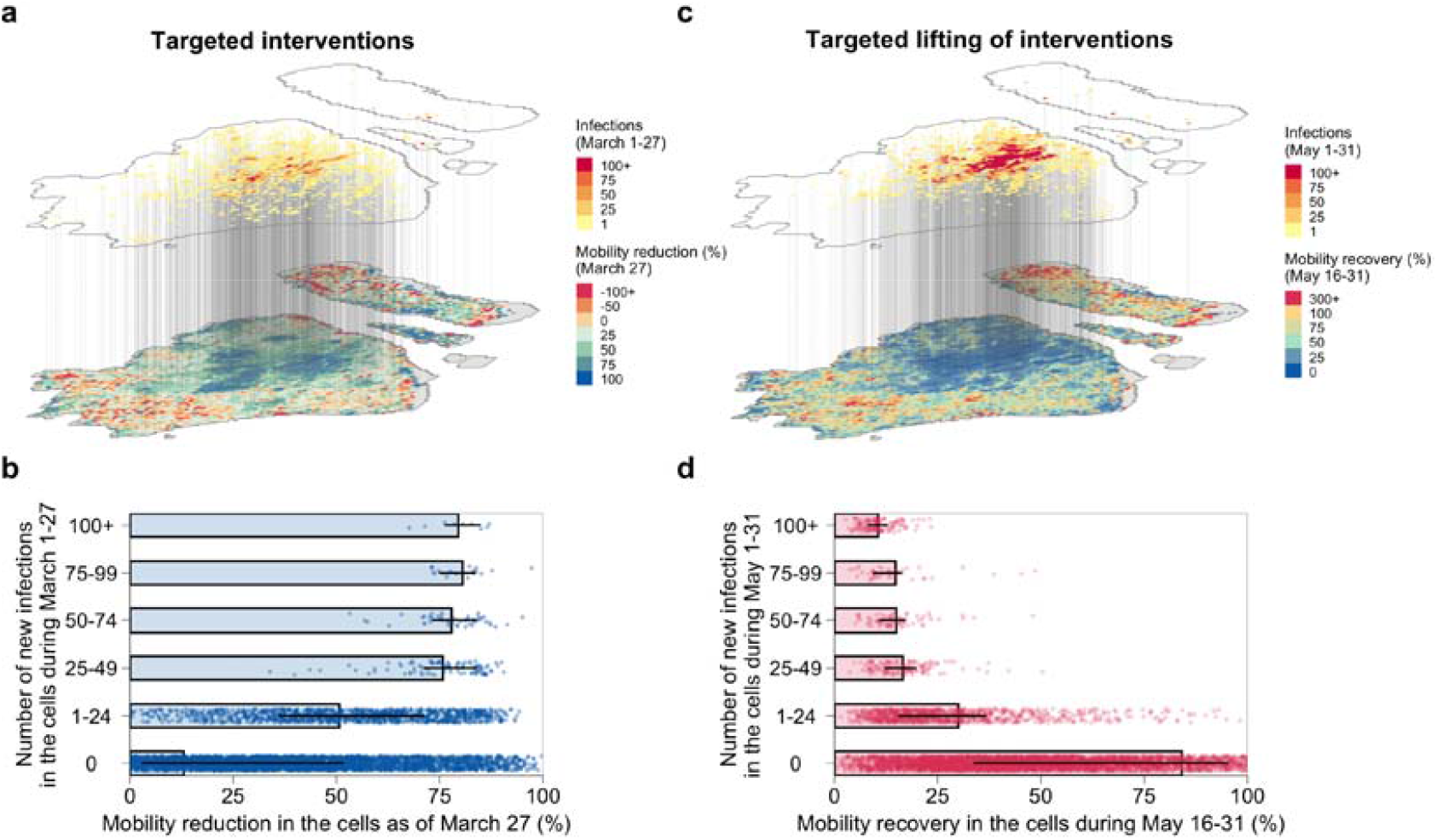
Impact of epidemic and interventions on the changes in mobility. **a**. The geographic distribution of infections and mobility reduction during the targeted interventions phase. The upper map represents the cumulative number of infections at the grid level as of March 27 (i.e., before the lockdown of eastern Shanghai). The lower map represents the mobility reduction, which is computed as the subtraction of daily trips on March 27 from the pre-outbreak mobility level, divided by the pre-outbreak mobility level. **b**. The mobility reduction as a function of number of new infections in the cells during the targeted interventions phase. The bar represents the mean value, while the horizontal line represents 50% quantile intervals. Each dot corresponds to the result for each cell. Note that the dots with a negative mobility reduction were not displayed. **c**. The geographic distribution of infections and mobility recovery during the targeted lifting of interventions phase. The upper map represents the cumulative number of infections at the grid level from May 1 to May 31. The lower map represents the mobility recovery, which is computed as the daily average trips between May 16 and May 31 divided by the pre-outbreak mobility level. The recovery may be beyond 100% if the daily trips during May 16 and May 31 are higher than the pre-outbreak mobility level. **d**. The same as panel b, but for the targeted lifting of interventions phase. Note that the dots with a mobility recovery beyond 100% were not displayed.

### Targeted interventions Phase

After the implementation of public places closures, school closures, mass screenings, and travel restrictions beginning on March 2, the number of daily trips decreased from 1.36 to 0.88 (see Fig. 1a). Long-distance trips, defined as those exceeding 30 km, experienced the most substantial decrease, dropping by approximately 47.5% compared to the pre-outbreak phase. This reduction brought the median travel distance down to 5.09 km (see Fig. 1c and d and SI Appendix, Fig. S4a). By the end of the targeted interventions phase, the number of communities within the mobility network had increased to around 50 (see Fig. 2b and SI Appendix, Fig. S3b and f).

### Citywide lockdown Phase

After a citywide lockdown was implemented on April 1, mobility decreased by 87.5% compared to the pre-outbreak phase and remained stable for about a month (see Fig. 1a). The median travel distance decreased to 1.21 km, with 79.0% of trips spanning less than 3 km (see Fig. 1c and d and SI Appendix, Fig. S4a). The initial 22 communities fragmented into 180 smaller ones, effectively dismantling the core-periphery structure that connected various parts of the city (see Fig. 2c and SI Appendix, Fig. S3c and f).

### Targeted lifting of interventions Phase

Coinciding with the partial lifting of interventions on May 1, data revealed a gradual increase in mobility, reaching 19.1% of pre-outbreak levels. Meanwhile, the median distance traveled per day rose to approximately half of what it was in the pre-outbreak phase (see Fig. 1a and c). The number of distinct communities decreased to 75 with a ramping up of the strength of connections across different regions of the city (see Fig. 2d and SI Appendix, Fig. S3d and f).

### Reopening Phase

Upon lifting most interventions on June 1, we observed an immediate resurgence in mobility flows, reaching 91.2% of pre-outbreak levels in under a week (see Fig. 1a). Short-distance trips (<3 km) increased more rapidly, exceeding pre-outbreak levels, while long-distance trips only recovered to about half of their pre-outbreak frequency. By June 30, the median trip distance had not returned to its level during targeted interventions (4.24 km vs. 5.09 km), although the number of daily trips had almost reverted to pre-outbreak figures (see Fig. 1c and SI Appendix, Fig. S4a). Ongoing mandatory COVID-19 tests for travel outside residential areas within the city, along with additional policies, prevented the community structure from fully reverting to its pre-outbreak state (43 vs. 22 communities) (see Fig. 2e and SI Appendix, Fig. S3e and f).

### Spatially heterogeneous impact of the epidemic and the adopted interventions

Before the lockdown of eastern Shanghai, mobility reductions were heterogeneous across regions, with larger reductions observed in regions severely hit by the epidemic (see Fig. 3a). Regions with more than 50 infections exhibited an average mobility reduction of 78.7%, while the reduction was just 13.0% for regions without infections (see Fig. 3b). During the targeted lifting of interventions phase, particularly after May 16 when public transportation began to resume, strict mobility-restricting policies persisted in high-risk areas with sustained incidence rates. In contrast, substantial rebounds in mobility were observed in low-risk regions, encompassing both suburban and rural areas of Shanghai (see Fig. 3c). Regions with more than 50 infections had a very low recovery of mobility (12.6% on average), while the recovery reached 84.1% for regions without infections (see Fig. 3d).

### Changes in frequency of travel, distance traveled, and mobility network community structure by demographic characteristics

To calculate the mobility and community structure by demographic characteristics, we analyzed mobility flows separately by age group and sex. The range of mobility was measured by the proportion of the area covered by the top ten communities (*α*), the total number of identified communities(*NC*), and the number of communities covering more than ten grid cells (*NC*_*g*≥10_). Based on the individual-level data of infected individuals reported between March 1 and March 25 (targeted interventions phase), we analyzed the relationship between mobility patterns and the incidence of SARS-CoV-2 as well as the number of cells with reported infections by demographic characteristics.

During the pre-outbreak phase, number of daily trips and distance travelled were highest for adults aged 30-59 years (6.20 km; 1.46 trips) and lowest for older adults aged 70+ (4.35 km; 0.60 trips) (see Fig. 4a and SI Appendix, Fig. S4b). Compared with middle-aged adults aged 30-59, individuals 0-18 years old travelled 38.2% less frequently and 23.6% shorter distances, and correspondingly had a 38.7% lower incidence and 58.0% less infected cells during the targeted interventions phase. For all age groups, higher mobility was correlated with higher infection incidence, and longer travel distances were correlated with larger infected areas (see SI Appendix, Table S7 and 8). Neither travel distance nor travel volume were obviously different across all age groups during the citywide lockdown; however, they quickly rebounded to the pre-outbreak level during the reopening phase (see Fig. 4b and c). Different age groups also presented significant differences in mobility network patterns across phases (see Fig. 4d-g). Middle-aged groups (30-59 years old) visited substantially more locations than younger or older groups; for example, the average degree ⟨*k*⟩ for middle-aged groups was 40 times that for 16-18 years old. Similarly, the mobility networks of middle-aged groups were more densely connected, with higher transitivity (adjacent neighboring locations) (see SI Appendix, Table S6). This difference was more prominent in community structures. For example, younger and older groups had smaller and less connected communities (*α*=5.93%, *NC*=319, *α* =6.88%, *NC*=369, respectively), whereas middle-aged groups had fewer well-connected communities covering large areas (*α =*37.25%, *NC*=142) (see SI Appendix, Fig. S5). The lockdown reduced the connection of the mobility networks for all age groups (see Fig. 4e).

**Figure 4.**
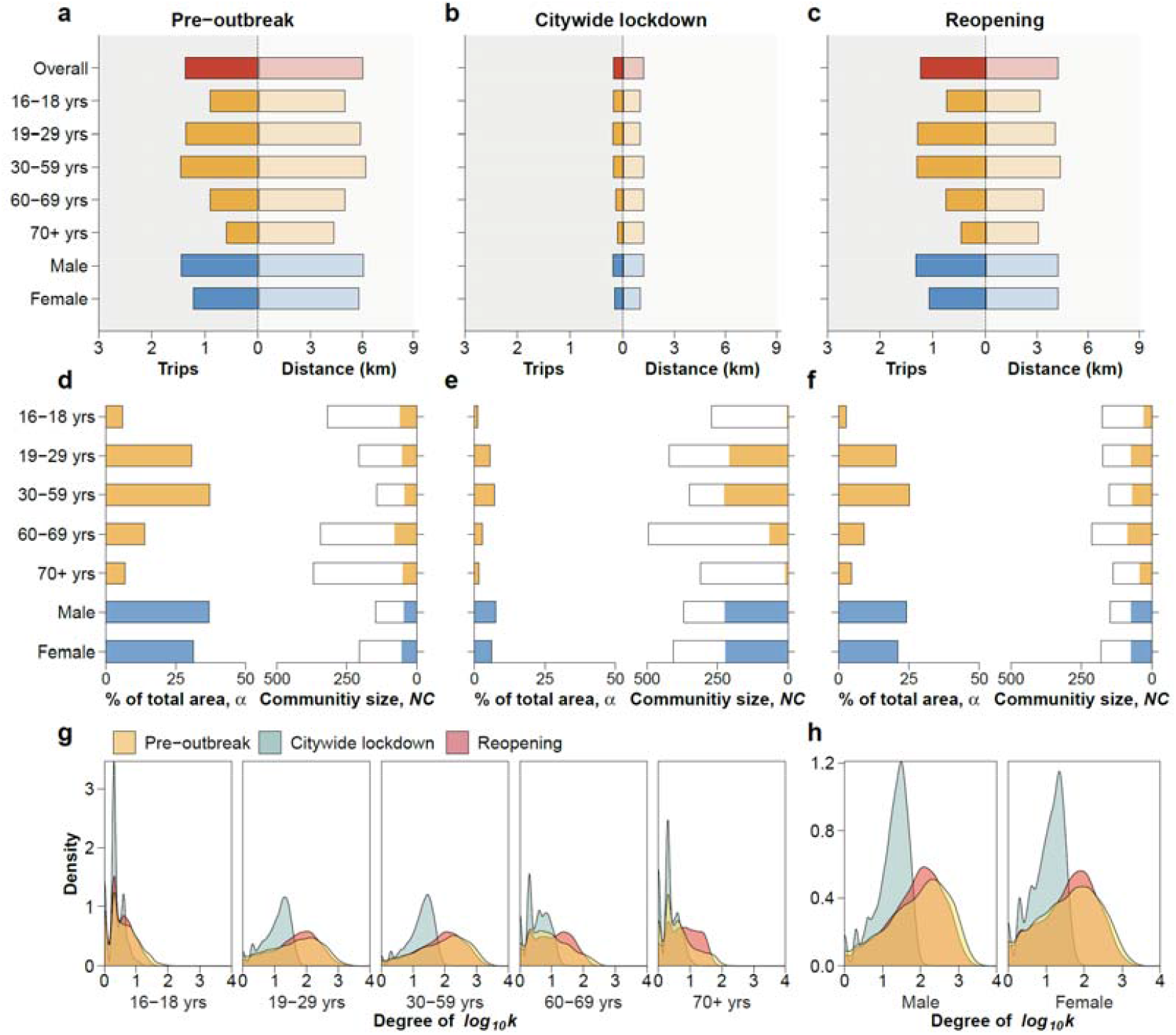
Changes in frequency, distance, and community structures of mobility network by age and sex. **a**-**c**. Mean number of daily trips and median distance travelled by age group and sex during the pre-outbreak, citywide lockdown, and reopening phases. Summary of frequency and distance across phases is shown in SI Appendix, Table S4-5. **d-f**. The left part of each panel represents the proportion, i.e., the top-10 communities in terms of area (1 km^2^) for each category to the total area (7,355 km^2^). The right part of each panel represents the number of identified communities. The filled portions represent the number of communities that spans more than 10 grids (*NC*_*g*≥10_), while the black box represents the overall number of communities (*NC*). **g**-**h**. The degree distribution of the mobility network across phases by age group and sex. Summary of the topological features of the mobility networks by age group and sex is shown in SI Appendix, Table S6.

Males were associated with longer travel distance (6.08 km vs. 5.81 km) and 30.6% higher daily trips than females during the pre-outbreak phase (see Fig. 4a and SI Appendix, Fig. S4c), which was associated with 9.7% higher incidence and 7.3% more infected cells than female during the targeted interventions phase (see SI Appendix, Table S7 and 8). There was no difference in mobility between males and females during citywide lockdown. The travel distance remained comparable across sexes (see Fig. 4b-c). Sex was also a strong factor affecting the mobility network patterns. During the pre-outbreak phase, males had a greater range of mobility and smaller community sizes (*α* =36.95%, *NC*=148) than females (*α* =31.37%, *NC*=205), indicating that males traveled more frequently and distantly than females. This difference persisted across all epidemic phases (see Fig. 4d-f and SI Appendix, Fig. S5).

### Additional analyses at different spatial and temporal resolutions

Additionally, we compared changes in frequency of daily trips at the grid, subdistrict, and district levels. Trips between subdistricts or districts exhibited higher reduction in mobility during the citywide lockdown for the subdistrict (91.3%) and district levels (95.4%) compared to the grid (1km×1km cells) level (87.5%) (see SI Appendix, Fig. S6a and SI Appendix, Table S9). We also observed a less marked reopening rebound of the mobility, reaching 79.1% and 69.2% of the pre-outbreak flows, respectively, for the subdistrict and district levels compared to 91.2% at the grid level. We then compared the proportion of daily population flows at different spatial resolutions, including inter-flow and intra-flow, where the inter-flow denotes the population flows between cells (or subdistricts, districts) and the intra-flow represents population flows within the same cell (or subdistrict, district). Our results show different patterns under different resolutions (see SI Appendix, Fig. S6b-d). Low-resolution mobility data may thus mask the variability in the dynamics of mobility flows.

We further investigated alterations in mobility and commuting patterns at various temporal resolutions. The periodic weekly commuting pattern swiftly rebounded during the reopening phase, even though the frequency of travel, distance traveled, and community structure had not fully recovered. Notably, we observed significant differences in travel frequency, distance traveled, and community structure of mobility networks between weekdays and weekends, as well as at different times of the day. For more details, refer to the SI Appendix Section 4 and SI Appendix, Fig. S7-9 for details.

## Discussion

Our analysis provided an in-depth assessment of the behavioral changes within the Shanghai population in response to the 2022 SARS-CoV-2 Omicron outbreak, considering fine spatial and temporal scales as well as demographic characteristics.

Pre-outbreak mobility was unevenly distributed across the city, with 33.4% of grids located in the center of Shanghai accounting for 80% of all trips. This is consistent with the geographical distribution of population density in Shanghai. The crowd movement during the pre-outbreak phase reveals the specific socio-economic distribution and commuting patterns in Shanghai. Mobility reductions were also spatially heterogeneous from the targeting interventions phase through the reopening phase, as different policies were adopted according to the local epidemic situation. Larger reductions were measured in regions more severely hit by the epidemic. These findings hint to possible spontaneous behavioral changes where individuals witnessing a large number of infections reported in their region might have limited their mobility beyond the mandated restrictions compared with those living in less affected regions. When the citywide lockdown entered into effect, the situation became homogenous as mobility reached its minimum level in all areas.

Throughout the outbreak, the frequency of travel and distance traveled generally adhered to the timeline of interventions implemented to combat the spread of SARS-CoV-2. Mobility reached its lowest level during the citywide lockdown phase, with an average of 0.17 trips per day and 1.21 km traveled. The community structure identified by the mobility flows followed the same pattern as well, with the population fragmenting into an increasing number of smaller communities as the level of intervention intensified. Mobility and community size quickly rebounded within the first week after interventions were lifted, although in the following month they had not fully recovered to pre-outbreak levels. During the outbreak, changes in behavior were spatially heterogenous within the city and directly associated with both the epidemic situation and interventions. We observed that males and individuals aged 30-59 years old traveled more frequently, traveled longer distances, and their communities were more connected, which were associated with higher incidence of SARS-CoV-2 infections and larger infected areas.

In late May, public transportation was partially reopened, and individuals living in less affected regions were allowed conditional trips (e.g., one individual per household per day was allowed to buy necessities). During the reopening phase, we found that mobility quickly rebounded within the first week (although it did not return to the pre-outbreak level). This recovering trend is substantially different from some European and US locations where the rebound was much slower, possibly due to the persistence of the epidemic or different levels of lockdown fatigue(12, 17, 21, 22). Within the Shanghai population, we found a slower mobility recovery during reopening among older adults (70+ years), which suggests possible spontaneous choices to limit mobility to minimize the risk of infection given widespread information about the increased risk of developing severe symptoms by age if infected. At the same time, it is also possible that the policy of requiring a negative PCR results within 72 hours to travel within the city (but outside their residential area) may have contributed to a reduced mobility among older adults as they are less likely to use smart phones to show proof of negative test result(23).

One interesting aspect of our analysis is that we have observed a spatially heterogeneous response to the outbreak. Although this was already found in previous country-level analyses(17, 24-26), here we are observing marked differences at the within-city scale. Our analysis is also showing that at the within-city scale, results are generally consistent if data is analyzed at 1 km^2^ resolution or using administrative boundaries (e.g., district, subdistrict), although quantitative differences to exists, highlighting the importance of selecting the appropriate spatial level of aggregation of mobility data depending on the focus research question. Moreover, we found that interventions altered not only the number of trips but also their length. In particular, after the lockdown was lifted, we observed an increase in trips under 3 km as compared to pre-outbreak mobility. These heterogeneous patterns may be useful for informing spatially targeted interventions at the within-city scale.

While mobile phone data is widely used to quantify human mobility, there are potential sources of inaccuracy to consider, such as i) population representativeness (e.g., by age), ii) geographical coverage, and iii) heterogeneity in user activity. First, our study may be subject to selection bias, as we analyzed the mobility of mobile phone owners, which could exclude or underrepresent young children and older adults (see SI Appendix, Fig. S1h and SI Appendix Section 1). However, despite this affecting our population-level results, we have provided an assessment by age and sex that does not suffer from this bias. Second, we analyzed data representing approximately 20% of Shanghai’s population, with a median coverage by subdistrict of 19.5% (interquartile range: 14.9%-24.7%) (see SI Appendix, Fig. S1c). Third, by relying on passively recorded cellular signaling data instead of actively recorded signals, we have mitigated the bias of heterogeneity in user activity. Another limitation is that the number of infections disaggregated by location, age, and sex is available to us only until March 25, 2022. This constraint limited our comparison between epidemiological data and human mobility patterns to the initial two phases of the outbreak.

In summary, behavioral changes during the 2022 Omicron outbreak were heterogeneous, both spatially and demographically. By shedding light on the varied responses among population groups, our findings can be instrumental in guiding the development of spatially targeted interventions to mitigate potential new surges in COVID-19 cases, as well as fostering preparedness for future respiratory infectious disease outbreaks.

## Materials and Methods

### Data sources

Mobile phone data. Cellular Signaling Data (CSD) were provided by China Unicom, one of the largest national mobile carriers in China, which accounts for approximately one-third of all active mobile phone users in Shanghai. Active signaling data was recorded during events such as phone calls, text messages, device power on/off, or tower switches, while passive signaling data captured the user’s location approximately every 30 minutes, provided the phone was turned on. The analyzed CSD data includes the timestamp of each event and a unique identifier for the mobile phone tower routing the activity. The data spans from February 15, 2022, to June 30, 2022, and consists of an average of 5.04 million phone users per day throughout the study period.

Epidemiological data. Daily aggregated data on the number of infections and individual-level data (line list) of all SARS-CoV-2 infections were extracted from multiple publicly available official data sources (websites of municipal health commission and local government media) as detailed in our previous study(3). The age and sex information are available only for infected individuals reported between March 1-March 25, 2022.

### Timeline of the outbreak and public health response

After the outbreak was initially reported on March 1, 2022, a series of non-pharmaceutical interventions (NPIs) were implemented to suppress transmission. Schools closed on March 12. From March 16 to 27, Shanghai introduced grid management by dividing subdistricts into high-risk and non-high-risk areas, based on factors such as the epidemiological situation (number of infections and cases), population density, social characteristics, and economic activity. High-risk areas underwent one or two rounds of population-wide PCR screening within 48 hours, accompanied by lockdown orders. Non-high-risk areas conducted a single round of mass screening.

On March 28, eastern Shanghai (comprising subdistricts east of the Huangpu River, see SI Appendix, Fig. S1a) entered a population-wide lockdown, followed by the rest of Shanghai on April 1 (citywide lockdown). Key enterprises and public transportation began resuming operations in May, with the citywide lockdown fully lifted on June 1. However, some restrictions persisted throughout June, limiting population movement. For instance, entering public places and transportation required proof of a negative PCR test result within 72 hours, and restaurants prohibited dine-in service until June 29. Additional details on the public health response can be found in SI Appendix, Fig. S2 and SI Appendix, Table S2.

### Definition of the five phases of the outbreak

For the purposes of this analysis, we categorized the outbreak into five phases based on the implemented interventions and the epidemic situation. The first phase, known as the “pre-outbreak phase,” spanned from February 15 to February 28, 2022. During this period, only a small number of sporadic and locally transmitted cases were recorded, and people’s daily activities remained largely unaffected. The period from February 1 to February 14 was excluded from our analysis as it is overlapped with the Chinese New Year holiday. The second phase is the “targeted interventions phase”, covering the period from March 1 to March 31, when spatially targeted NPIs were deployed to suppress transmission. The third phase is the “citywide lockdown phase”, covering the period from April 1 to April 30, when the entire city was in lockdown. The fourth phase is the “targeted lifting of interventions phase”, covering the period from May 1 to May 31, when restrictions started to gradually scale-down in specific areas of the city. The last phase is the “reopening phase”, covering the period from June 1 to June 30, when policies started to be lifted throughout the entire city.

### Frequency and distance of daily trips

A trip was counted when a user switched to one or more new cell towers, until the user became stationary again (no further switch for approximately 30 min). We only consider trips between different cells of the grid. We defined as *T*_*ij*_ (*t*) the number of trips between grid *j* and grid *i* at time *t*. The average number of trips per individual at time *t* was thus defined as ⟨*T*⟩ (t) = T(t)/*U*(t), where T(t) = ∑_*i* ≠ *j*_ *T*_*ij*_ (t) represents the total number of trips at time *t*, and *U*(*t*) represents the number of active users at time *t* (which dynamically changes over time due to the flow of commuter to and from Shanghai). To quantify to what extent mobility changed during the outbreak, we compare the mobility during different epidemic phases to a baseline phase with pre-outbreak mobility. Estimates were disaggregated by age, sex, day type (i.e., weekday and weekend), and time of the day (i.e., daytime and nighttime).

### Definition of the mobility network and community detection

To investigate structural changes in the mobility network throughout various stages of the outbreak, we reconstructed the mobility network *G*_*P*_ for each phase *P*. In this network, each node represents a cell of the grid, with directed edges connecting nodes where users move between cell *i* and cell *j*. The degree of node *i* is then defined by 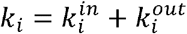, where 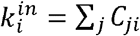, and 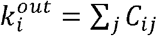, where *C*_*ji*_ indicates whether node *j* is connected to node *i* or not (i.e., users travel from node *j* to node *i*). The average degree ⟨*k*⟩ is then calculated as 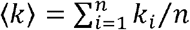, where *n* is the number of nodes. The number of days in each phase *P* is denoted by *D*_*P*_. Subsequently, the edge weights *w*_*ij*_(*P*) are calculated as the average daily number of trips between cells during this phase as 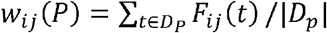. We exclude edges whose average weight is below the threshold *w*_*ij*_(*D*) < 1.

We used the Infomap method(20) to detect the community structures in the mobility network. Briefly, considering the sequence of communities visited by a random walker who will tend to linger within communities, the algorithm detects the community based on the probability distribution of random walks. A community partition is regarded as good if the description of that sequence requires relatively little information, in the sense of Shannon entropy, and the Infomap method is built to optimize the minimum description length of the random walk on the network. Compared with other methods, this approach retains the information about the directions and weights of the edges, which has the advantage of being flexible for finding community structures on large weighted and directed networks(27, 28). To assess community detection, we calculate the modularity(29). As an index of the difference of connectivity within a community versus between-communities, a relatively lower modularity value indicates a higher strength of connections between different communities rather than within the same community.

The same methods were used to analyze the mobility networks and community structures by demographic characteristics by subsetting the dataset to consider only mobility flows for the analyzed population group.

### Ethical Considerations

This study was approved by the institutional review board of the School of Public Health, Fudan University (IRB# 2022-05-0969).

## Data Availability

Mobile phone data are proprietary and confidential. We obtained access to these data from the SmartSteps company controlled by China Unicom within the framework of the COVID-19 research project. To safeguard the privacy of the users, CSD was aggregated over time and space scale and by users'age group and sex. Raw mobility data cannot be made publicly available to preserve privacy. Grid-level data to reproduce the findings of this study can be requested from the corresponding author.

## Data Availability

Mobile phone data are proprietary and confidential. We obtained access to these data from the SmartSteps company controlled by China Unicom within the framework of the COVID-19 research project. To safeguard the privacy of the users, CSD was aggregated over time and space scale and by users’ age group and sex(30). Raw mobility data cannot be made publicly available to preserve privacy. Grid-level data to reproduce the findings of this study can be requested from the corresponding author.

## Code availability

The code will be made available on GitHub upon acceptance of the manuscript.

## Acknowledgments

H.Y. acknowledges financial support from the Key Program of the National Natural Science Foundation of China (No. 82130093). X.L. is supported by the National Nature Science Foundation of China (No. 72025405, 72001211, 82041020, 72088101), National Social Science Foundation of China (No. 22ZDA102), and the Hunan Science and Technology Plan Project (No. 2020TP1013). J.Z. is supported by the Shanghai Rising-Star Program (No. 22QA1402300).

## Notes

### Competing Interest Statement

The authors have declared no competing interest.

