## Supplementary Materials for "Heterogeneous changes in mobility in response to the SARS-CoV-2 Omicron BA.2 outbreak in Shanghai"

### Supplementary section 1. Data description

#### Data representativeness

We compared the demographic characteristics of China Unicom users to all mobile phone users and the 2020 census population in Shanghai(1) to determine whether mobile data is representative of the general population.

China Unicom had an average of 5.04 million users (range: 4.62-5.42 million) in Shanghai during the study period (SI Appendix, Fig. S1e), accounting for about 27% of all mobile phone users in Shanghai. Demographic characteristics (i.e., age and sex) of the study population remained constant over time (SI Appendix, Fig. S1f and g). Since demographic data is unavailable for all mobile phone users in Shanghai, we inferred these estimates based on the smartphone penetration rate from the Statistical Report on Internet Development in mainland China(2). We found that a higher proportion of adults aged 20-59 and males were included in the study population compared to all mobile phone users. This bias is also present when comparing the study population to the Shanghai census population in 2020 (SI Appendix, Fig. S1h, Table S1).

Additionally, our analyzed users correspond to about 20% of Shanghai population with a median coverage by subdistrict of 19.5% (interquartile range: 14.9%-24.7%) (SI Appendix, Fig. S1c).

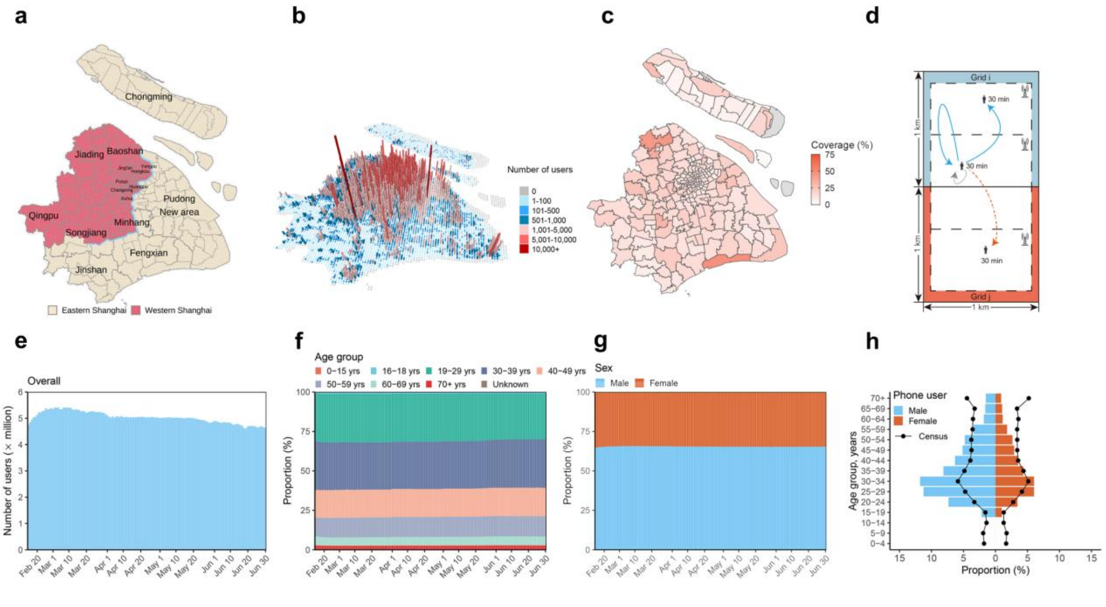

**Supplemental Figure 1. Geographical and demographical distribution of analyzed users in Shanghai.**

**a.** Geographic division of Shanghai. Shanghai is divided into 16 districts (dark gray) and 216 subdistricts (gray), then further divided into 7,355 1km×1km grids (light gray). Eastern and western Shanghai are separated by the Huangpu River (blue layer). **b.** The number of analyzed users living in each cell. Each user's home location was assumed to be the cell with the longest visit duration between 12:00 AM and 6:00 AM per month. We only considered regularly active users that visited their home location at least 15 days per month. **c.** Geographical coverage of analyzed users at the scale of subdistrict. **d.** An illustration of how mobility is recorded. A trip is counted when a user switches to one or more new cell towers, until the user becomes stationary again (no further switch for approximately 30 min). Trips can be within the same cell (blue, solid line) or between different cells (red, dotted-dashed line). We only considered the trips between different cells in this study. Movements without changing cell towers are not recorded (gray, dotted line). **e.** Daily changes in the number of analyzed users over time. **f-g.** Proportion of analyzed users by age group and sex. **h.** Proportion of study population by age and sex. Black dotted lines denote expected distributions based on age-sex distributions derived from census 2020.

**Supplementary Table 1. Comparison between analyzed users, inferred total mobile phone users, and 2020 census population in Shanghai.**

| Characteristics | Analyzed users (%) | Mobile phone users (%) * | Census population (%) |
|---|---|---|---|
| **Total** | 5,037,355 (100) | 18,254,754 (100) | 24,870,895 (100) |
| **Age** | | | |
| 0-19 yrs | 50,381 (1.00) | 1,737,438 (9.52) | 3,146,480 (12.65) |
| 20-29 yrs | 1,522,875 (30.23) | 5,004,717 (27.42) | 3,719,990 (14.96) |
| 30-39 yrs | 1,519,713 (30.17) | 3,855,543 (21.12) | 5,037,103 (20.25) |
| 40-49 yrs | 897,459 (17.82) | 3,010,053 (16.49) | 3,633,785 (14.61) |
| 50-59 yrs | 636,528 (12.64) | 2,568,730 (14.07) | 3,518,075 (14.15) |
| 60+ yrs | 392,359 (7.79) | 2,078,274 (11.38) | 5,815,462 (23.38) |
| Unknown | 18,040 (0.36) | - | - |
| **Sex** | | | |
| Male | 3,295,604 (65.42) | 9,401,198 (51.50) | 12,875,211 (51.77) |
| Female | 1,741,749 (34.58) | 8,853,556 (48.50) | 11,995,684 (48.23) |

Note: * Mobile phone users in Shanghai were inferred based on the smartphone penetration rate from Statistical Report on Internet Development in China(2).

#### Data processing

First, we analyzed the spatial resolution of the CSD data at the tower level (up to 150 meters in the central urban areas of Shanghai) by aggregating the data into 7,355 grids of equally sized 1km×1km cells. Then, we analyzed the data at the administrative division level, where the grids were further aggregated into the subdistrict or district levels (SI Appendix, Fig. S1a). For grids that span multiple subdistricts, the flows within the grid were distributed to the subdistricts in proportion to the area occupied by the grid on different subdistricts.

We examined demographic characteristics by extracting the age and sex of mobile phone users according to their registration information.

#### Timeline of NPIs

The public health response and data sources in details were reported in SI Appendix, Fig. S2 and Table S2.

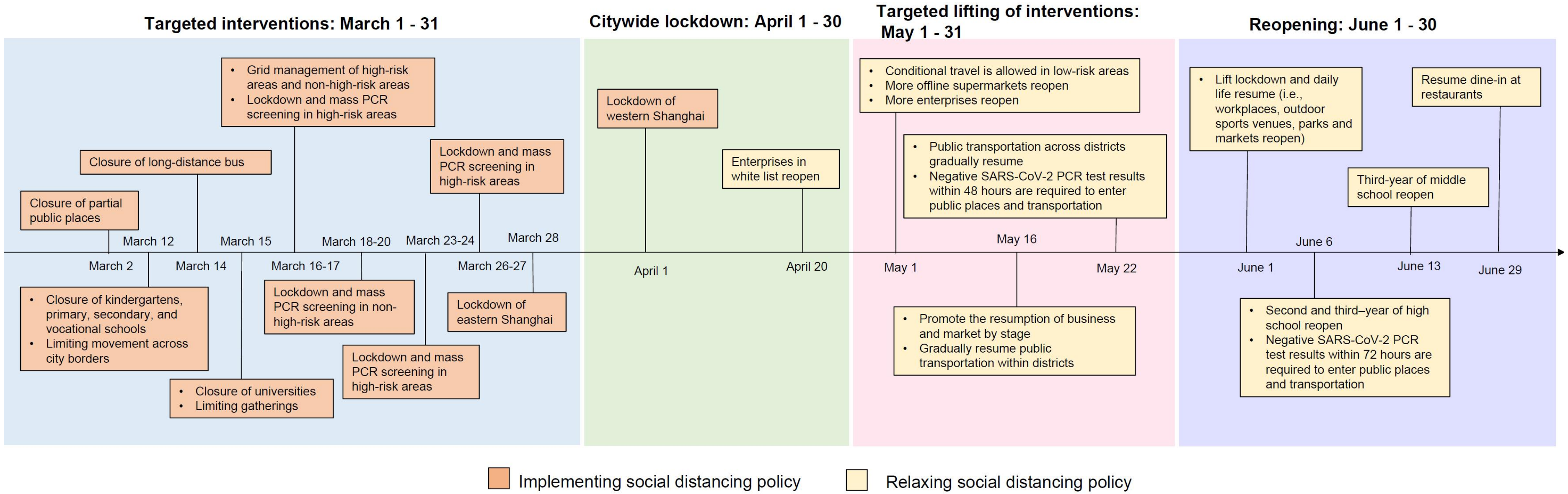

**Supplemental Figure 2. Timeline of the public health response in Shanghai across phases.**
More details on the public health response are provided in the Supplementary Table 2.

**Supplementary Table 2. The timeline of implementing and relaxing social distancing policies in Shanghai.**

| Interventions | Time | link |
|---|---|---|
| **Implementing social distancing policy** | | |
| Closure of partial public places | 2022/3/2 | http://www.xinwenmh.com/38679.html |
| Closure of kindergartens, primary, secondary, and vocational schools | 2022/3/12 | https://mp.weixin.qq.com/s/_yiS-7IhSbp6J9vmQQBYDQ |
| Limiting movement across city borders | 2022/3/12 | https://mp.weixin.qq.com/s/Kev1hnuiRxEqSwjY5J8lpw |
| Closure of long-distance bus | 2022/3/14 | https://mp.weixin.qq.com/s/dku0LLKZUf1hidcQcjCEZQ |
| Closure of universities | 2022/3/15 | https://mp.weixin.qq.com/s/_yiS-7IhSbp6J9vmQQBYDQ |
| Limiting gatherings | 2022/3/15 | https://baijiahao.baidu.com/s?id=1727354297326963152&wfr=spider&for=pc |
| Grid management of high-risk areas and non-high-risk areas | 2022/3/16-3/27 | https://mp.weixin.qq.com/s/ZJSpoQYjGupSBOsWtSr_vA |
| Lockdown and mass PCR screening in high-risk areas | 2022/3/16-3/17 | https://mp.weixin.qq.com/s/ZJSpoQYjGupSBOsWtSr_vA |
| Lockdown and mass PCR screening in non-high-risk areas | 2022/3/18-3/20 | https://mp.weixin.qq.com/s/r6OBkpYSHRezqzHXXVclkw |
| Lockdown and mass PCR screening in high-risk areas | 2022/3/23-3/24 | https://mp.weixin.qq.com/s/VU5d7WNiv7HF6DASXCaYNQ |
| Lockdown and mass PCR screening in high-risk areas | 2022/3/26-3/27 | https://mp.weixin.qq.com/s/_OKOPwbPfOw1Zi48FOvF4w |
| Lockdown of eastern Shanghai | 2022/3/28 | https://mp.weixin.qq.com/s/Ufza89hhBGZsiGPTHoC5aQ |
| Lockdown of western Shanghai | 2022/4/1 | https://mp.weixin.qq.com/s/Ufza89hhBGZsiGPTHoC5aQ |
| **Relaxing social distancing policy** | | |
| Enterprises in white list reopen | 2022/4/20 | https://baijiahao.baidu.com/s?id=1730429764028680040&wfr=spider&for=pc |
| Conditional travel is allowed in low-risk areas | 2022/5/1 | https://mp.weixin.qq.com/s/5vJr5Nc4E_UwTw8znAL8Nw |
| More offline supermarkets reopen | 2022/5/1 | https://mp.weixin.qq.com/s/Uu1z33046PUktCm9DZml4g |
| More enterprises reopen | 2022/5/1 | https://mp.weixin.qq.com/s/2ti_8CvS-nBIxNCEcTfXNg |
| Promote the resumption of business and market by stage | 2022/5/16 | https://mp.weixin.qq.com/s/F7F6wll914R7Zpn6hYd78g |
| Gradually resume public transportation within districts | 2022/5/16 | https://www.thepaper.cn/newsDetail_forward_18124628 |
| Transportation to outside Shanghai gradually resume | 2022/5/16 | https://mp.weixin.qq.com/s/YInymvwy4HOobWKqxOuJJw |

| | | |
|---|---|---|
| Public transportation cross districts gradually resume, and negative SARS-CoV-2 PCR test results within 48 hours are required to enter public places and transportation | 2022/5/22 | https://mp.weixin.qq.com/s/l1FsMY8Dg3l4XJAJDJnxAA |
| Lift lockdown and daily life resume. For example, workplaces, outdoor sports venues, parks and markets reopen. | 2022/6/1 | https://mp.weixin.qq.com/s/4dZErfcWAFp6V5bdAfajbg |
| Second and third–year of high school reopen | 2022/6/6 | https://mp.weixin.qq.com/s/PGwmoFMMjgH03ABt6ys64w |
| Negative SARS-CoV-2 PCR test results within 72 hours are required to enter public places and transportation | 2022/6/6 | https://sghexport.shobserver.com/html/baijiahao/2022/06/06/762433.html |
| Third-year of middle school reopen | 2022/6/13 | https://mp.weixin.qq.com/s/PGwmoFMMjgH03ABt6ys64w |
| Resume dine-in at restaurants | 2022/6/29 | https://mp.weixin.qq.com/s/5urWguFkYAIdirZDXYDqYg |

**Supplementary section 2. Additional results of mobility and community structure**

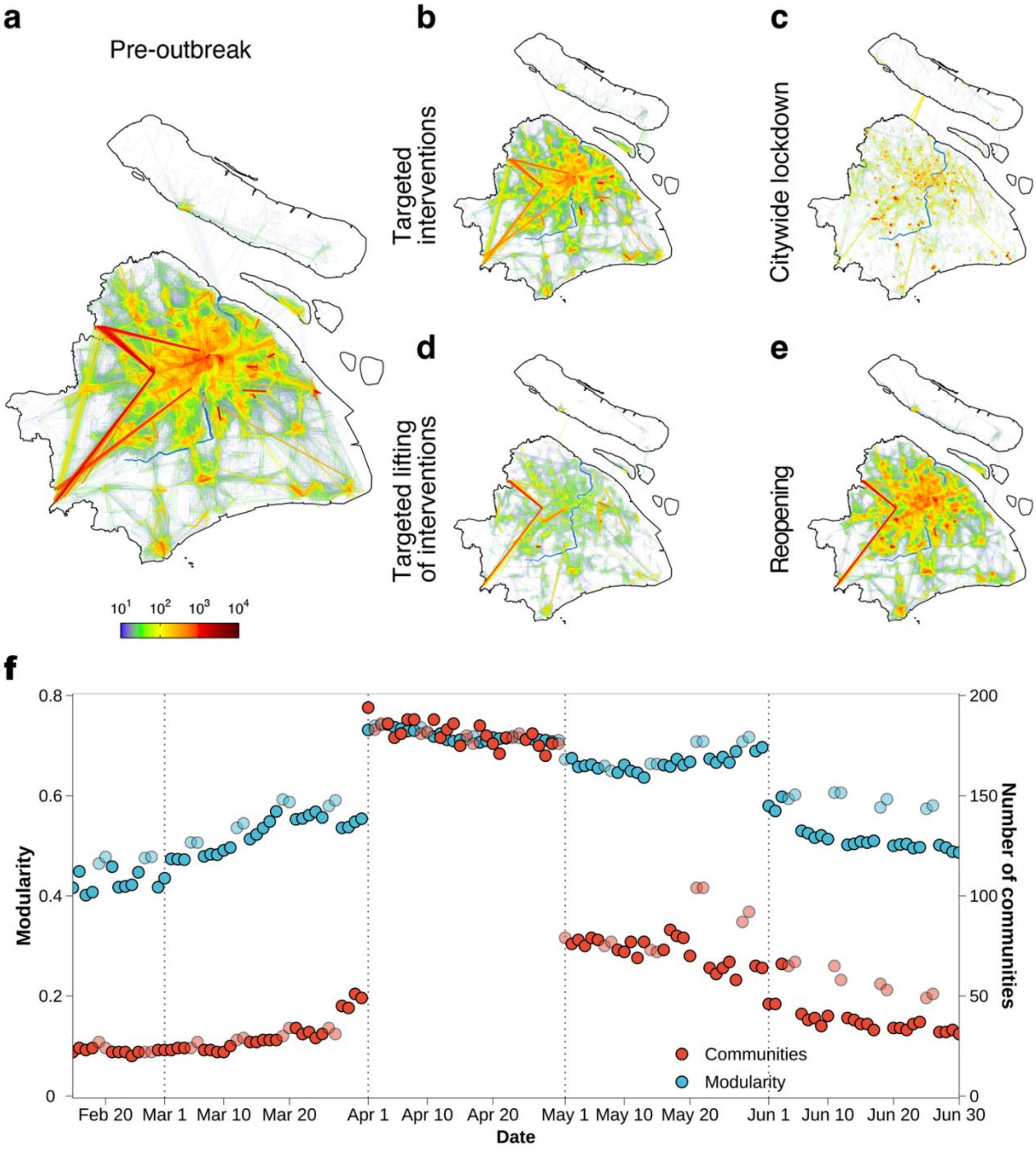

**Supplemental Figure 3. Spatial mobility network across phases.**
**a.** A depiction of the networks during the pre-outbreak phase. Line color and width indicate the number of daily trips on average along each connection. The blue line represents the Huangpu River, separating eastern Shanghai and western Shanghai. Trips below 10 were omitted from the data. **b-e.** Same as a. but during the targeted interventions, citywide lockdown, targeted lifting of interventions, and reopening phases. **f.** The time series of modularity (blue) and the number of identified communities (red) across phases. Light and dark

colors correspond to weekends and weekdays, respectively. The variation of modularity is associated with NPIs' stringency across phases. It increased drastically from about 0.4 during the pre-outbreak phase to about 0.7 during the citywide lockdown, and then gradually decreased with the relaxation of NPIs stringency. Summary of the topological features of mobility networks across phases is shown in Supplementary Table 3.

**Supplementary Table 3. Summary of the basic topological features of the mobility networks across phases.**

Nodes (*n*) represent the grid visited, and edges (*m*) represent the trip from grid *i* to grid *j*, respectively. $\langle k \rangle$ is the average degree. *C* and *r* are the clustering coefficient(3) and the assortative coefficient(4), respectively. *E* is the network efficiency.

| Network | *n* | *m* | $\langle k \rangle$ | *C* | *r* | *E* |
|---|---|---|---|---|---|---|
| **Pre-outbreak** | 6164 | 1016055 | 330.640 | 0.571 | 0.172 | 0.400 |
| **Targeted interventions** | 6074 | 712061 | 234.462 | 0.561 | 0.282 | 0.357 |
| **Citywide lockdown** | 6118 | 94334 | 30.838 | 0.537 | 0.354 | 0.203 |
| **Targeted lifting of interventions** | 6029 | 210530 | 69.839 | 0.555 | 0.184 | 0.267 |
| **Reopening** | 6170 | 769238 | 249.348 | 0.631 | 0.283 | 0.354 |

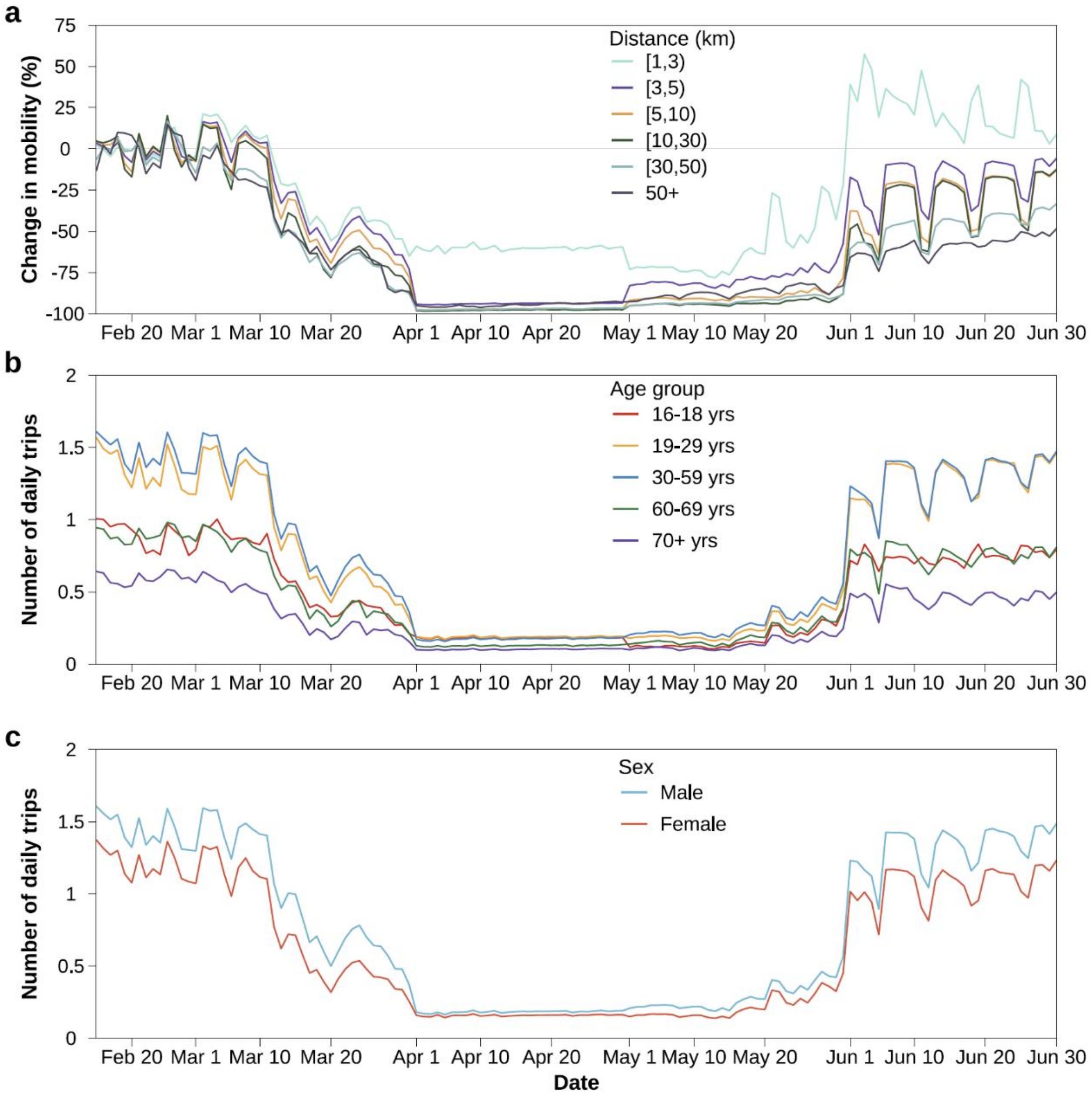

**Supplemental Figure 4. Relative mobility changes in different distance ranges and number of daily trips by age group and sex over time.**

The number of daily trips during the pe-outbreak phase (February 15-28) were set to 0 as a baseline to estimate the relative mobility changes over time.

**Supplementary Table 4. Frequency of daily trips across phases.**

| | **Pre-outbreak** Mean (95%CI) | **Targeted interventions** Mean (95%CI) | **Citywide lockdown** Mean (95%CI) | **Targeted lifting of interventions** Mean (95%CI) | **Reopening** Mean (95%CI) |
|---|---|---|---|---|---|
| **Overall** | 1.36 (1.23, 1.52) | 0.88 (0.40, 1.50) | 0.17 (0.16, 0.18) | 0.26 (0.17, 0.46) | 1.24 (0.93, 1.39) |
| **Age** | | | | | |
| 16-18 yrs | 0.90 (0.75, 1.00) | 0.58 (0.25, 0.97) | 0.18 (0.17, 0.19) | 0.17 (0.11, 0.33) | 0.74 (0.66, 0.83) |
| 19-29 yrs | 1.36 (1.19, 1.55) | 0.87 (0.39, 1.51) | 0.19 (0.18, 0.20) | 0.25 (0.16, 0.44) | 1.28 (0.96, 1.45) |
| 30-59 yrs | 1.46 (1.32, 1.61) | 0.95 (0.44, 1.59) | 0.18 (0.16, 0.18) | 0.29 (0.19, 0.49) | 1.30 (0.98, 1.46) |
| 60-69 yrs | 0.90 (0.83, 0.98) | 0.55 (0.25, 0.95) | 0.13 (0.12, 0.13) | 0.20 (0.12, 0.35) | 0.75 (0.58, 0.85) |
| 70+ yrs | 0.60 (0.53, 0.65) | 0.36 (0.17, 0.62) | 0.10 (0.10, 0.11) | 0.14 (0.09, 0.23) | 0.46 (0.35, 0.54) |
| **Sex** | | | | | |
| Male | 1.45 (1.31, 1.60) | 0.96 (0.45, 1.58) | 0.18 (0.17, 0.19) | 0.29 (0.19, 0.49) | 1.32 (1.00, 1.48) |
| Female | 1.21 (1.08, 1.37) | 0.73 (0.30, 1.33) | 0.16 (0.15, 0.17) | 0.22 (0.14, 0.40) | 1.07 (0.79, 1.21) |
| **Day type** | | | | | |
| Weekday | 1.39 (1.24, 1.52) | 0.92 (0.38, 1.50) | 0.17 (0.16, 0.18) | 0.26 (0.17, 0.45) | 1.30 (1.12, 1.39) |
| Weekend | 1.29 (1.24, 1.39) | 0.79 (0.45, 1.28) | 0.17 (0.16, 0.18) | 0.28 (0.18, 0.43) | 1.06 (0.86, 1.19) |
| **Time of the day** | | | | | |
| Daytime | 1.05 (0.91, 1.12) | 0.65 (0.30, 1.09) | 0.10 (0.09, 0.11) | 0.19 (0.11, 0.35) | 0.94 (0.71, 1.05) |
| Nighttime | 0.32 (0.19, 0.43) | 0.23 (0.10, 0.41) | 0.07 (0.07, 0.08) | 0.07 (0.06, 0.12) | 0.29 (0.21, 0.34) |

Note: 95%CI denote the 2.5% and 97.5% quantile.

**Supplementary Table 5. Distance of daily trips across phases.**

| | **Pre-outbreak** Median (IQR) | **Targeted interventions** Median (IQR) | **Citywide lockdown** Median (IQR) | **Targeted lifting of interventions** Median (IQR) | **Reopening** Median (IQR) |
|---|---|---|---|---|---|
| **Overall** | 6.04 (2.99, 13.03) | 5.09 (2.50, 11.29) | 1.21 (1.00, 2.24) | 2.83 (1.38, 5.06) | 4.24 (2.06, 10.11) |
| **Age** | | | | | |
| 16-18 yrs | 5.00 (2.24, 12.05) | 4.00 (2.15, 8.75) | 1.00 (1.00, 2.07) | 2.24 (1.28, 4.99) | 3.16 (1.49, 7.28) |
| 19-29 yrs | 5.91 (2.92, 12.98) | 5.00 (2.24, 10.91) | 1.00 (1.00, 2.23) | 2.83 (1.34, 5.00) | 4.06 (2.00, 9.88) |
| 30-59 yrs | 6.20 (3.01, 13.38) | 5.24 (2.82, 11.67) | 1.21 (1.00, 2.24) | 2.91 (1.40, 5.11) | 4.35 (2.11, 10.61) |
| 60-69 yrs | 4.98 (2.24, 10.21) | 4.12 (2.21, 8.54) | 1.21 (1.00, 2.24) | 2.24 (1.31, 4.23) | 3.38 (1.99, 7.79) |
| 70+ yrs | 4.35 (2.23, 9.49) | 3.60 (2.08, 7.80) | 1.21 (1.00, 2.24) | 2.24 (1.29, 4.11) | 3.08 (1.59, 7.05) |
| **Sex** | | | | | |
| Male | 6.08 (2.99, 13.56) | 5.11 (2.67, 11.67) | 1.21 (1.00, 2.24) | 2.91 (1.41, 5.12) | 4.24 (2.06, 10.30) |
| Female | 5.81 (2.94, 12.05) | 5.00 (2.24, 10.3) | 1.00 (1.00, 2.23) | 2.53 (1.31, 4.48) | 4.24 (2.05, 9.76) |
| **Day type** | | | | | |
| Weekday | 6.08 (3.00, 13.11) | 5.24 (2.82, 11.41) | 1.21 (1.00, 2.24) | 3.00 (1.75, 5.37) | 4.46 (2.23, 10.78) |
| Weekend | 5.81 (2.83, 12.79) | 4.98 (2.24, 10.44) | 1.21 (1.00, 2.24) | 2.24 (1.13, 4.28) | 3.08 (1.25, 8.02) |
| **Time of the day** | | | | | |
| Daytime | 6.08 (3.00, 13.10) | 5.11 (2.75, 11.37) | 1.00 (1.00, 2.24) | 2.83 (1.32, 5.00) | 4.24 (2.06, 10.20) |
| Nighttime | 5.81 (2.85, 12.83) | 5.00 (2.24, 11.05) | 1.21 (1.00, 2.24) | 3.00 (1.85, 5.38) | 4.12 (2.07, 10.01) |

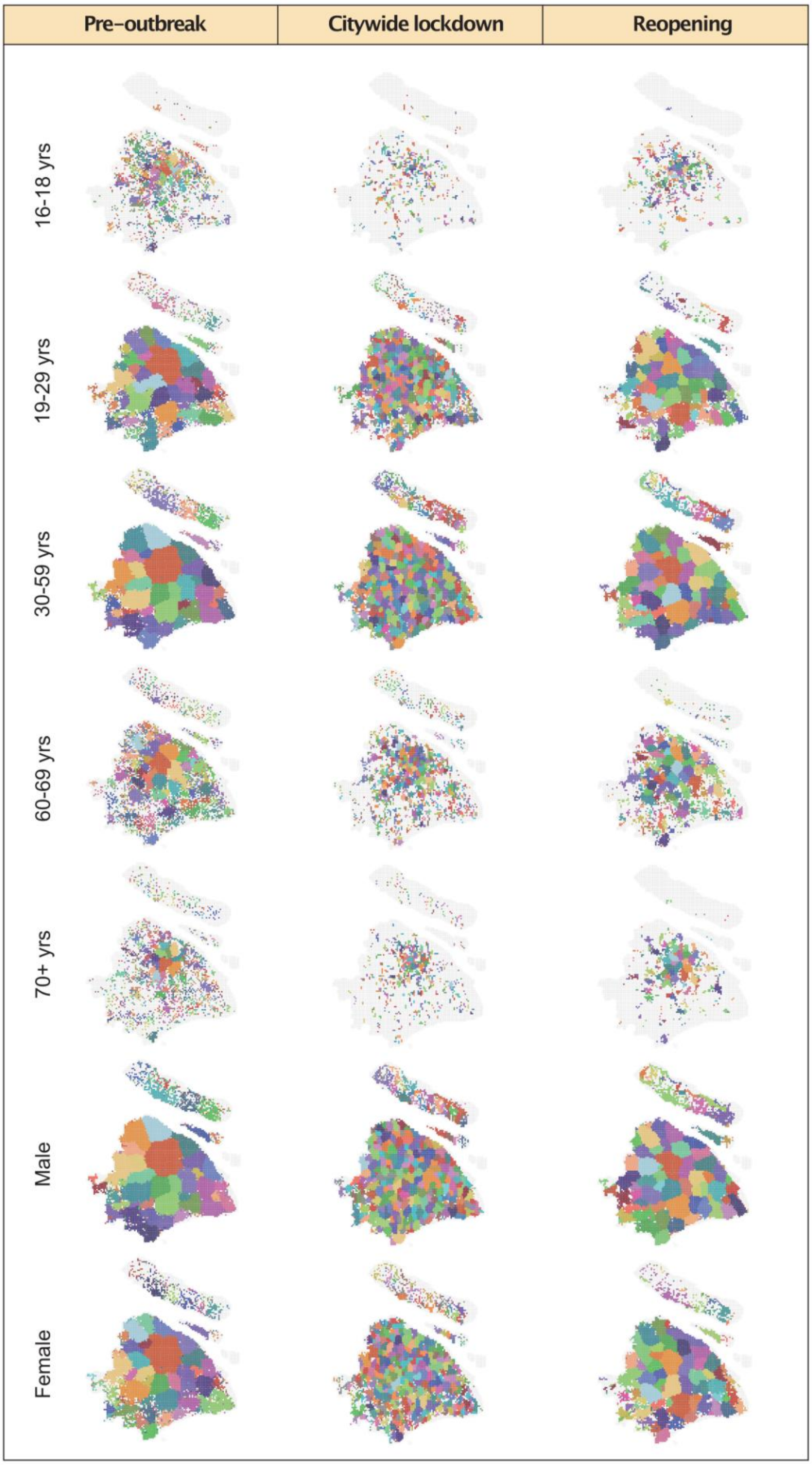

**Supplemental Figure 5. Distribution of the communities by age group and sex for pre-outbreak, citywide lockdown, and reopening phases.** The color of the community partition corresponds to the size of the community.

**Supplementary Table 6. The basic topological features of the mobility networks by age group, sex, day type, and the time of day.**

Nodes ($n$) represent the grid visited, and edges ($m$) represent the trip from grid $i$ to grid $j$, respectively. $\langle k \rangle$ is the average degree. $C$ and $r$ are the clustering coefficient(3) and the assortative coefficient(4), respectively. $E$ is the network efficiency.

| Network | $n$ | $m$ | $\langle k \rangle$ | $C$ | $r$ | $E$ |
|---|---|---|---|---|---|---|
| **Pre-outbreak** | | | | | | |
| 16-18 yrs | 2067 | 6663 | 6.447 | 0.118 | 0.242 | 0.125 |
| 19-29 yrs | 5223 | 385710 | 147.697 | 0.501 | 0.123 | 0.353 |
| 30-59 yrs | 5934 | 754282 | 254.224 | 0.547 | 0.208 | 0.381 |
| 60-69 yrs | 3960 | 48233 | 24.36 | 0.271 | 0.45 | 0.198 |
| 70+ yrs | 2282 | 10845 | 9.505 | 0.17 | 0.423 | 0.093 |
| Male | 5990 | 796775 | 226.035 | 0.551 | 0.194 | 0.385 |
| Female | 5409 | 395167 | 146.115 | 0.503 | 0.155 | 0.341 |
| Weekday | 6210 | 1060715 | 341.615 | 0.539 | 0.169 | 0.403 |
| Weekend | 6393 | 1089779 | 340.929 | 0.531 | 0.164 | 0.413 |
| Daytime | 6014 | 835057 | 277.704 | 0.574 | 0.183 | 0.389 |
| Nighttime | 5212 | 299115 | 114.779 | 0.487 | 0.218 | 0.339 |
| **Citywide lockdown** | | | | | | |
| 16-18 yrs | 768 | 1040 | 2.708 | 0.134 | 0.479 | 0.003 |
| 19-29 yrs | 4938 | 39760 | 16.104 | 0.442 | 0.315 | 0.136 |
| 30-59 yrs | 5904 | 69469 | 23.533 | 0.511 | 0.331 | 0.181 |
| 60-69 yrs | 2513 | 6101 | 4.855 | 0.204 | 0.346 | 0.019 |
| 70+ yrs | 1005 | 1401 | 2.788 | 0.102 | 0.301 | 0.004 |
| Male | 5973 | 75517 | 25.286 | 0.515 | 0.345 | 0.186 |
| Female | 5199 | 40234 | 15.478 | 0.443 | 0.343 | 0.127 |
| Weekday | 6125 | 95512 | 31.188 | 0.536 | 0.351 | 0.208 |
| Weekend | 6178 | 97745 | 31.643 | 0.531 | 0.347 | 0.212 |
| Daytime | 5836 | 63715 | 21.835 | 0.496 | 0.371 | 0.176 |
| Nighttime | 5599 | 54468 | 19.456 | 0.485 | 0.312 | 0.153 |
| **Reopening** | | | | | | |
| 16-18 yrs | 1032 | 2487 | 4.82 | 0.235 | 0.044 | 0.029 |
| 19-29 yrs | 4963 | 290622 | 117.115 | 0.587 | 0.242 | 0.314 |
| 30-59 yrs | 5953 | 572704 | 192.409 | 0.609 | 0.312 | 0.333 |
| 60-69 yrs | 3091 | 34105 | 22.067 | 0.444 | 0.553 | 0.164 |
| 70+ yrs | 1251 | 6651 | 10.633 | 0.396 | 0.36 | 0.076 |
| Male | 5999 | 594707 | 198.269 | 0.616 | 0.303 | 0.338 |
| Female | 5197 | 303298 | 116.72 | 0.584 | 0.262 | 0.3 |
| Weekday | 6163 | 835396 | 271.1 | 0.624 | 0.267 | 0.362 |
| Weekend | 6295 | 618394 | 196.471 | 0.598 | 0.295 | 0.346 |
| Daytime | 5979 | 626217 | 209.472 | 0.627 | 0.284 | 0.34 |
| Nighttime | 5270 | 232182 | 88.115 | 0.576 | 0.313 | 0.296 |

**Supplementary Table 7. Travel frequency and reported SARS-CoV-2 infections by age and sex between March 1 and March 25, 2022**

| Characteristics | Number of reported infections (n=9,851) | Incidence of reported infections (per 1,000) | Relative difference in incidence (%) | Relative difference in travel frequency (%) |
|---|---|---|---|---|
| **Age** | | | | |
| 0-18* yrs | 936 (9.5) | 0.297 | -38.7 | -38.2 |
| 19-29 yrs | 1,381 (14.0) | 0.371 | -23.5 | -7.5 |
| 30-59 yrs | 5,916 (60.1) | 0.485 | Ref | Ref |
| 60-69 yrs | 969 (9.8) | 0.284 | -41.5 | -42.2 |
| 70+ yrs | 649 (6.6) | 0.270 | -44.3 | -62.9 |
| **Sex** | | | | |
| Male | 5,326 (54.1) | 0.414 | 9.7 | 30.6 |
| Female | 4,525 (45.9) | 0.377 | Ref | Ref |

Note: We only analyzed the mobility data for the age group of 16-18 years old.

**Supplementary Table 8. Distance travelled and number of cells with reported infections by age and sex between March 1 and March 25, 2022**

| Characteristics | Number of cells with reported infections (n=7,355) | Relative difference in number of cells (%) | Relative difference in distance (%) |
|---|---|---|---|
| **Age** | | | |
| 0-18* yrs | 462 | -58.0 | -23.6 |
| 19-29 yrs | 598 | -45.6 | -4.7 |
| 30-59 yrs | 1,099 | Ref | Ref |
| 60-69 yrs | 446 | -59.4 | -21.3 |
| 70+ yrs | 312 | -71.6 | -31.2 |
| **Sex** | | | |
| Male | 1,053 | 7.3 | 2.2 |
| Female | 981 | Ref | Ref |

Note: We only analyzed the mobility data for the age group of 16-18 years old.

### Supplementary section 3. Additional analyses at different spatial resolutions

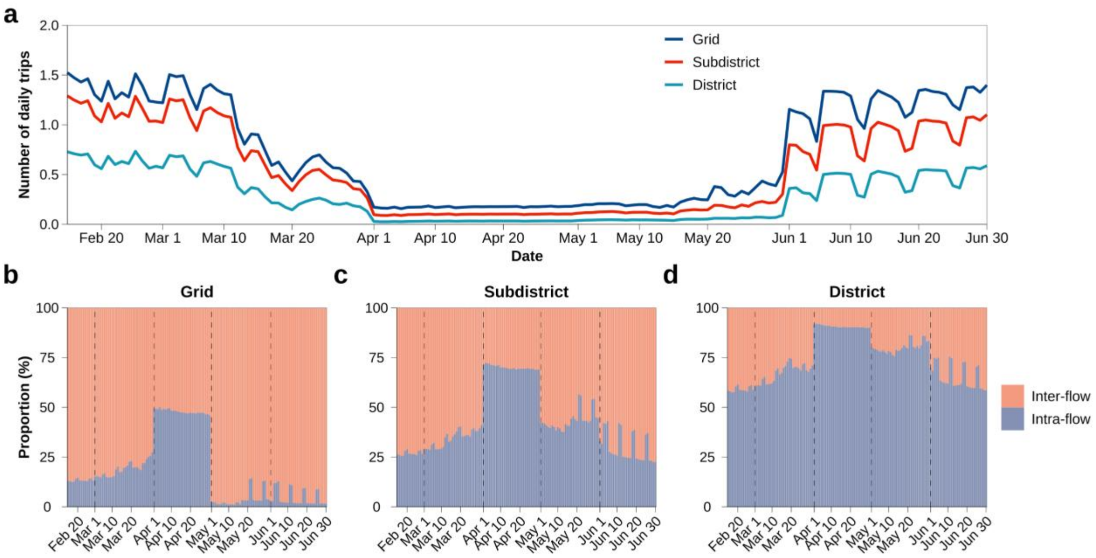

**Supplemental Figure 6. Changes in population flows at different spatial resolutions.**

**a.** Changes in number of daily trips per individual over time. The blue, red, and green lines represent the daily trips aggregated at the grid, subdistrict, and district level, respectively. **b-d.** Proportion of daily population flows at grid, subdistrict, and district level, respectively.

**Supplementary Table 9. Frequency of daily trips across phases at different spatial resolutions.**

| | **Grid level** Mean (95%CI) | **Subdistrict level** Mean (95%CI) | **District level** Mean (95%CI) |
|---|---|---|---|
| **Pre-outbreak** | 1.36 (1.23, 1.52) | 1.15 (1.03, 1.29) | 0.65 (0.56, 0.73) |
| **Targeted interventions** | 0.88 (0.40, 1.50) | 0.72 (0.32, 1.25) | 0.37 (0.14, 0.69) |
| **Citywide lockdown** | 0.17 (0.16, 0.18) | 0.10 (0.09, 0.10) | 0.03 (0.03, 0.03) |
| **Targeted lifting of interventions** | 0.26 (0.17, 0.46) | 0.15 (0.11, 0.25) | 0.05 (0.04, 0.08) |
| **Reopening** | 1.24 (0.93, 1.39) | 0.91 (0.61, 1.09) | 0.45 (0.26, 0.58) |

Note: 95%CI denote the 2.5% and 97.5% quantile.

### Supplementary section 4. Additional analyses at different temporal resolutions

#### Hourly travel geographic and temporal variability

The hourly population movements provide a high-resolution dynamical characterization of how the population reacted to the COVID-19 outbreak. Prior to the targeted interventions, full lockdowns, or district-based controls, human mobility exhibits a distinct commuting pattern during the workday: the travel volume during rush hours (7-9 AM, 5-7 PM) accounted for 47.2% of the daily trips on weekdays, most of which were work-related trips. Weekend crowd movements are instead mostly driven by recreational trips, with travel volume peaking around 4 PM and extending from late morning to early evening. The time series of travel volume exhibits a strong weekly periodic pattern in the pre-outbreak phase and the early outbreak stage in March 2022. We observed a dramatic drop in mobility across Shanghai as a result of the citywide lockdown on April 1. The most effective reduction in overall mobility occurred during rush hours (91.3%) and was associated with a disruption of commuting patterns. Moreover, the trips reduced to the bare minimum (0.2 daily trips on average, about one-eighth of the pre-outbreak phase) that is required to maintain essential living and working conditions.  During the targeted lifting of interventions phase, trips gradually rebounded as a result of the partial reopening. The mobility has materially improved during the reopening phase and returned to its 7-days periodicity in June, with the relaxation of restricting policies (SI Appendix, Fig. S7a).

To further investigate the commuting pattern, we analyzed the spatial heterogeneity of netflow (inflow minus outflow) during rush hours. We defined *commercial-* or *residential-dominant* cells in terms of the pre-outbreak phase (the *commercial-dominant cell* is with netflow>25 during the morning rush hours, while the *residential-dominant cell* is with netflow<-25). Approximately, *commercial-dominant* zones accounted for 17.8% of the 7,355 grids and were primarily located in the central urban areas of Shanghai, while *residential-dominant* zones accounted for 28.1% of grids and were located in adjacent suburban and rural areas (SI Appendix, Fig. S7b). Despite being dispersed unevenly throughout the district, they were highly symmetrical during morning and evening rush hours. This distinction of mobility patterns between *commercial-dominant* and *residential-dominant* zones gradually waned in March, then disappeared in the citywide lockdown and targeted lifting of interventions phases (SI Appendix, Fig. S8).

To assess changes in mobility, we compared the number of daily trips per individual at the hourly scale in February between the pre-outbreak phase and subsequent phases (SI Appendix, Fig. S7c). The largest reduction in average hourly population mobility occurred in April (87.2%) and May (80.6%) with respect to the pre-outbreak phase. While the periodic weekly pattern quickly rebounded in the reopening phase, the hourly travel trips reached over 90% of the pre-outbreak level, indicating that Omicron's impact on the Shanghai population had yet to vanish during the reopening phase.

**Frequency of travel, distance travelled, and community structure by day type and time of the day**

We explored the frequency, distance travelled, and community structure of mobility networks by day type and time of the day (SI Appendix, Fig. S9).

During the pre-outbreak phase, there is no obvious difference in distance and daily trips between weekday and weekend, which remained during the targeted interventions and citywide lockdown phases. However, shorter distance and a smaller number of daily trips were observed on weekends during reopening, indicating that the social activities had yet to return to normal. Moreover, all phases, except for the citywide lockdown phase, showed little difference in distance between daytime and nighttime trips; although, number of daily trips was 3 times higher during the daytime than the nighttime.

There is a noticeable difference between population movements on weekdays ($\alpha$=40.97%) and weekends ($\alpha$=24.12%) during the pre-outbreak phase. Additionally, in contrast to the relatively quick recovery of weekday traffic during the reopening phase, recreational weekend trips rebounded slowly and remained closer to home. This can be seen at the subdistrict level where intra-subdistrict flow accounted for 40% of weekend travel volumes in the reopening phase compared to 28% of weekend trips during the pre-outbreak phase. We also found that the number of identified communities on weekends is larger than that on weekdays in the reopening phase (SI Appendix, Fig. S3f). Likewise, more locations were visited during the daytime ($\alpha$=36.32%) than the nighttime ($\alpha$=24.87%), and the average degree $\langle k \rangle$ for the daytime was two times that of the nighttime. However, the mobility difference o essentially disappeared between daytime ($\langle k \rangle$=23.89) and nighttime ($\langle k \rangle$=19.45) during the reopening phase, when quarantine impact was felt on a wider population- or geographic-level.

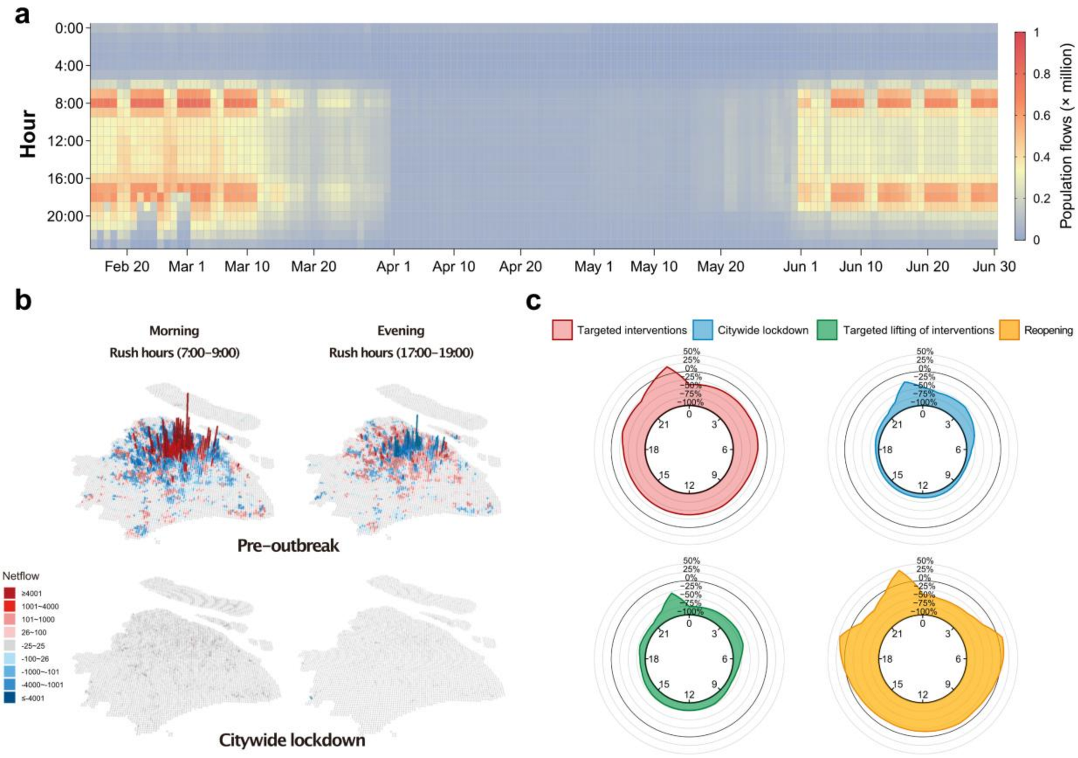

**Supplemental Figure 7. Hourly mobility changes in Shanghai.**

**a.** Hourly population movements from February 15 to June 30, 2022. Warmer colors indicate more trips and cooler colors represent less trips. **b.** The geographic distribution of the grid-level netflow (inflow minus outflow) during weekday rush hours throughout the pre-outbreak phase and citywide lockdown, respectively. Red and blue bars represent positive and negative netflow, which we called *commercial-* or *residential-dominant* cells on the corresponding locations. The cell is defined as *commercial-dominant* if its netflow>25 during the morning rush hours, while defined as *residential-dominant* if its netflow<-25). **c.** The hourly change in mobility compared with the pre-outbreak phase. The change ratio is computed as the average hourly trips per individual over the targeted interventions, citywide lockdown, targeted lifting of interventions, and reopening phases, with respect to the trips during the pre-outbreak phase.

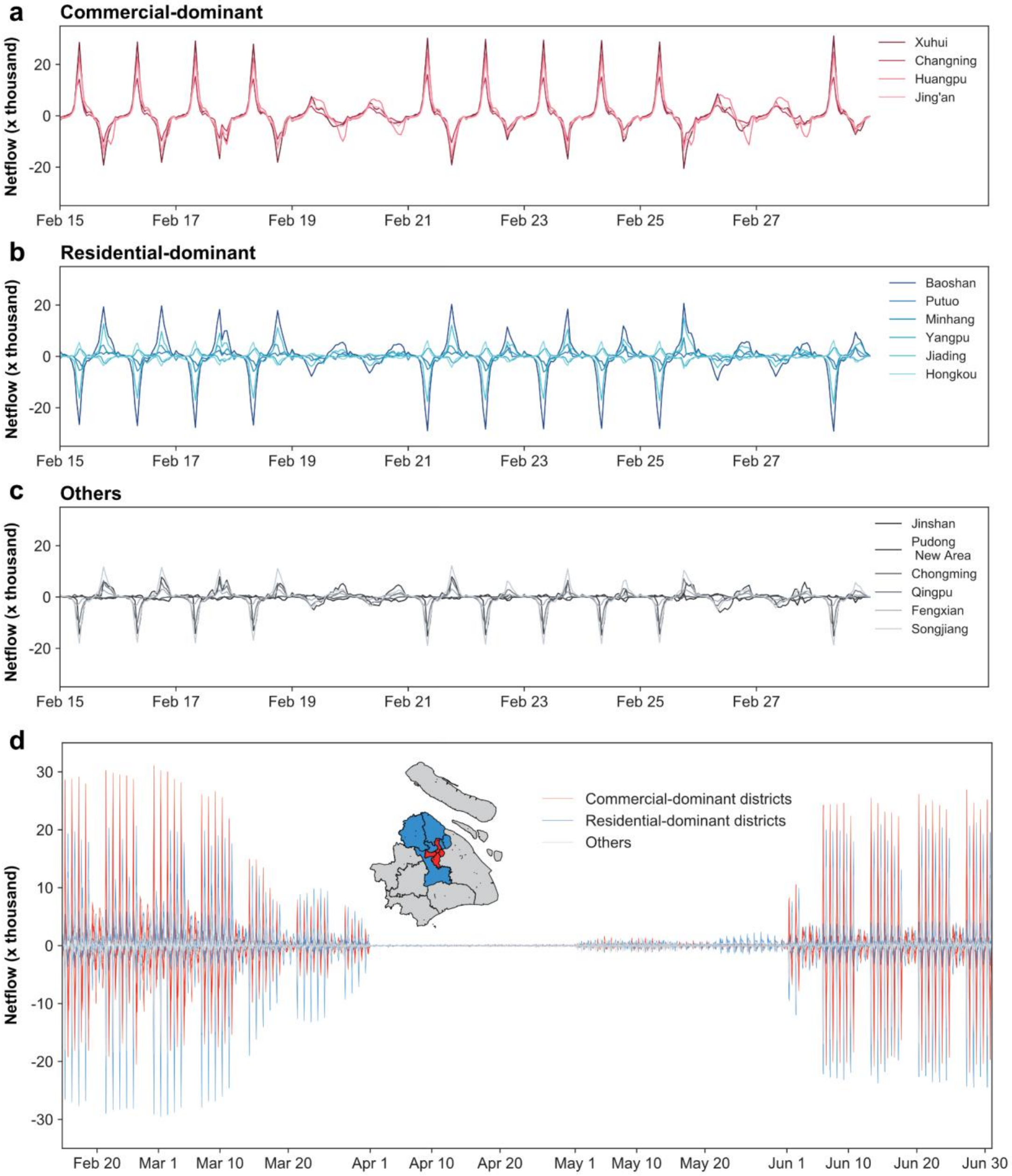

**Supplemental Figure 8. District-level netflow fluctuation across phases. a-c.** Three types of districts throughout the pre-outbreak phase. The classification results are as follows: *Commercial -dominant* districts: Xuhui, Changning, Huangpu, Jing'an; *Residential-dominant* districts: Baoshan, Hongkou, Minhang, Jiading, Putuo, Yangpu; Others: Pudong New Area, Fengxian, Jinshan, Songjiang, Qingpu, Chongming. **d**. District-level netflow fluctuated across phases. We divided Shanghai's 16 districts into categories based on the grid types that compose the majority of each district. Red and blue lines represent *commercial*- and *residential-dominant* districts, respectively.

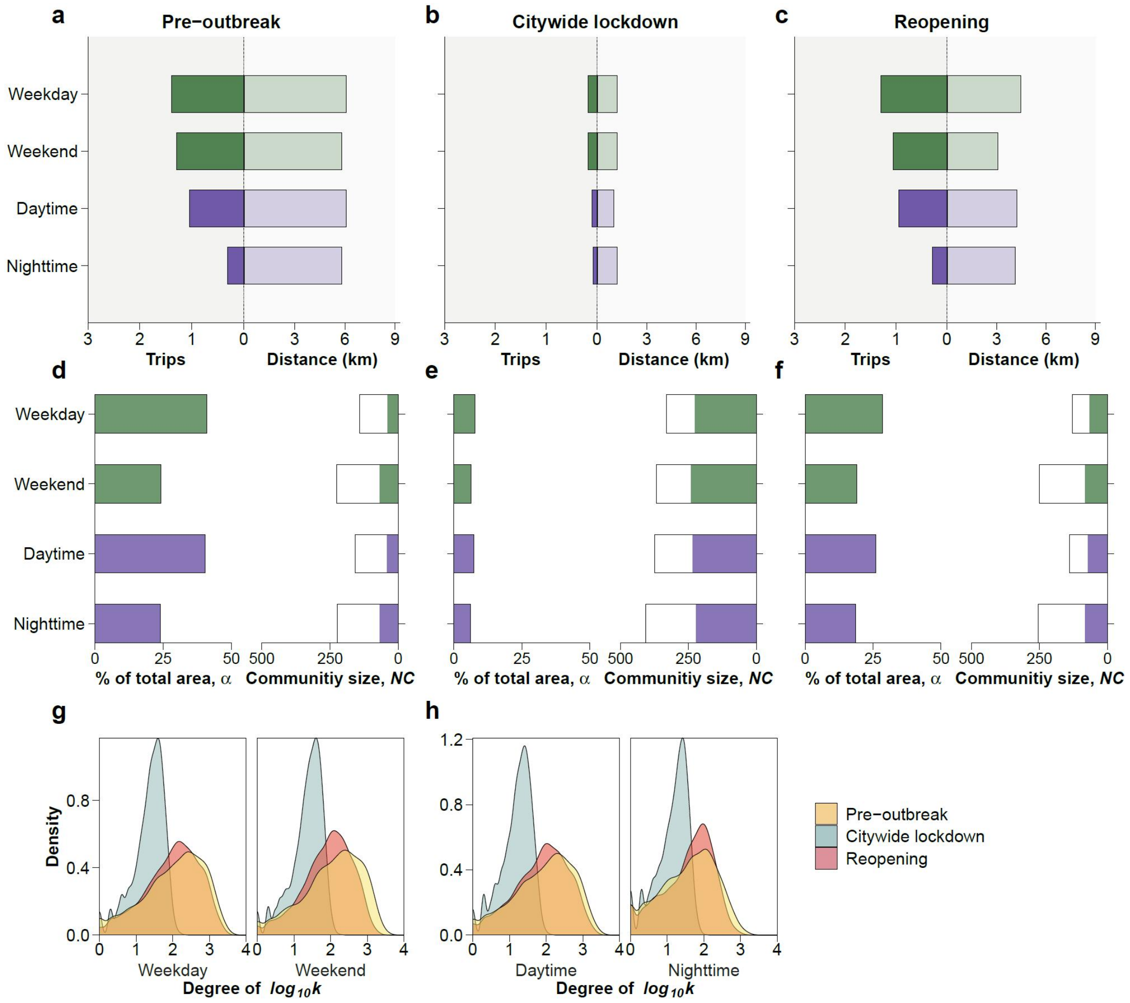

**Supplemental Figure 9. Changes in frequency, distance, and community structures of mobility network by day type and time of the day.**
**a-c.** Median distance of daily trips and number of daily trips by day type and time of the day during the pre-outbreak, citywide lockdown, and reopening phases. The summary of frequency and distance travelled across phases is shown in Supplementary Tables 4-5. **d-f.** The left part of each panel represents the proportion $\alpha$, i.e., the top-10 communities in terms of area (1 km$^2$) for each category to the total area (7,355 km$^2$). The right part of each panel represents the number of identified communities, and those filled suggest the number of communities that covered more than 10 grids $NC_{g\geq 10}$, while the black box indicates the overall community size $NC$. **g-h.** Degree distribution of the mobility network across phases by data type and time of the day. Summary of the topological features of the mobility networks by day and time of the day is shown in Supplementary Table 6.
